# Indexing Healthcare Access and Quality for Surgically Amenable Causes of Death: A Global Analysis of 204 Countries and Territories from 1990 to 2019

**DOI:** 10.1101/2024.02.04.24302290

**Authors:** Siddhesh Zadey, Emily R. Smith, Catherine A. Staton, Tamara N. Fitzgerald, Joao Ricardo Nickenig Vissoci

## Abstract

**Background:** We analyzed the healthcare access and quality (HAQ) index for surgically amenable causes, its progress over time, and the gap compared to non-surgical HAQ across 204 countries and territories from 1990 to 2019 for children (up to 14 years) and overall populations.

**Study Design:** The Global Burden of Disease 2019 study provided mortality-to-incidence ratios and risk-standardized death rates for 32 causes with preventable mortality. Of these, 14 (18) and 9 (17) causes were considered surgical (non-surgical) for overall population and children, respectively. We constructed composite indices ranging from 0 (worst) to 100 (best) using the adjusted Mazziotta Pareto index methodology. The ratio of surgical HAQ in 2019 to that in 1990 noted change over time. Surgical-to-non-surgical HAQ ratio gave the relative gap in 2019. Ratios >1 depicted improvement over time or better performing surgical care systems.

**Results:** In 2019, the overall surgical HAQ varied from 18.00 for the Central African Republic to 98.25 for Canada. The child surgical HAQ index varied from 39.87 for Chad to 99.41 for San Marino. For both surgical HAQ indices, 202 countries noted progress from 1990 to 2019. Only 31 countries (15.2%) had greater surgical HAQ index values than their non-surgical counterparts. The child surgical HAQ index lagged behind non-surgical for 61.28% of countries.

**Conclusion:** Low-income countries had limited progress in surgical HAQ indices since 1990 and lagged behind non-surgical HAQ index in 2019 the most. These findings are valuable for global evaluations, policymaking, and advocacy for investing in surgical care.

## Introduction

Health systems exist to prevent population-level mortality and morbidity. The Healthcare Access and Quality (HAQ) index using the Global Burden of Disease (GBD) study is a composite metric of 32 causes of preventable mortality for comparison across regions and progress tracking over time.(1, 2) The HAQ index is adapted from the amenable mortality due to healthcare framework in response to the World Health Report 2000 for better international comparison of health systems.(3) The most recent country-wise estimates for the overall HAQ index are now available for 2019.(2) These include an overall HAQ index (ages 0–74 years) and those for select age groups: the young (ages 0–14 years), working (ages 15–64 years), and post-working (ages 65–74 years) groups.

HAQ specific to surgical care has previously been estimated using data until 2016.(4) Of the 32 causes, there are 14 that are amenable to surgery, and thus, mortality risk could be reduced by involving surgeons, anesthetists, or obstetricians. While the Lancet Commission of Global Surgery (LCoGS) indicators assess surgical preparedness, delivery, and impact, HAQ index assesses surgical care systems’ impact on population health outcomes. These composite indices are crucial for comparing monitoring surgical care systems, policymaking, and resource allocation. However, updated surgical HAQ index estimates are missing. About 40% of the 5 billion people lacking access to timely, safe, and affordable surgical care are children.(5) Yet, focus on pediatric surgery in policymaking has been limited.(6) In part, this is due to lack of measures that can help assess the amenable mortality due to pediatric surgical care. A surgical HAQ index for children can fill the assessment gap. However, such an index is missing.

We had the following aims: 1. To systematically analyze the HAQ indices for surgically amenable causes for overall population and children (0-14 years) across 204 countries and territories from 1990 to 2019. 2. To track the relative changes in the overall and child surgical HAQ indices during this period for countries and across the World Bank country income groups. 3. To compare the overall and child surgical HAQ indices relative to the corresponding non-surgical HAQ indices.

## Methods

We obtained HAQ data from the Global Burden of Disease (GBD) 2019 Study.(2) Briefly, the GBD Study uses data from health facility administrative records, censuses, clinical trials, demographic surveillance, disease registries, environmental monitoring such as satellite data, epidemiological surveillance including disease outbreaks notifications, surveys, and vital registration across countries. These data are then cleaned and collated and used as inputs for several statistical models that provide estimates for prevalence, mortality, disability-adjusted life-years, and several other outcomes of interest. The annual mean estimates with 95% uncertainty intervals are modeled for 204 countries/territories across three decades. GBD focuses on the face validity/internal consistency of the estimates.(7) The validity and reliability of estimates is also demonstrated by the out-of-sample prediction metrics. Generally, estimates for recent years can be thought to be more precise than those for earlier years due to the increase in data density for several LMICs. This is also reflected in the tighter uncertainty intervals in recent years. More details can be found in the source studies.(8, 9) The HAQ index values are based on estimates for mortality-to-incidence ratios and risk-standardized death rates from the GBD.

Of the 32 HAQ index causes, 14 were considered surgically amenable.(4) For children up to 14 years of age, IHME models 26 causes of death for the HAQ, leaving out those that do not pertain to this age group. We considered nine as surgically amenable. **Table 1** provides the list of causes considered for surgical and non-surgical HAQ indices for overall and child populations.

**Table 1:**
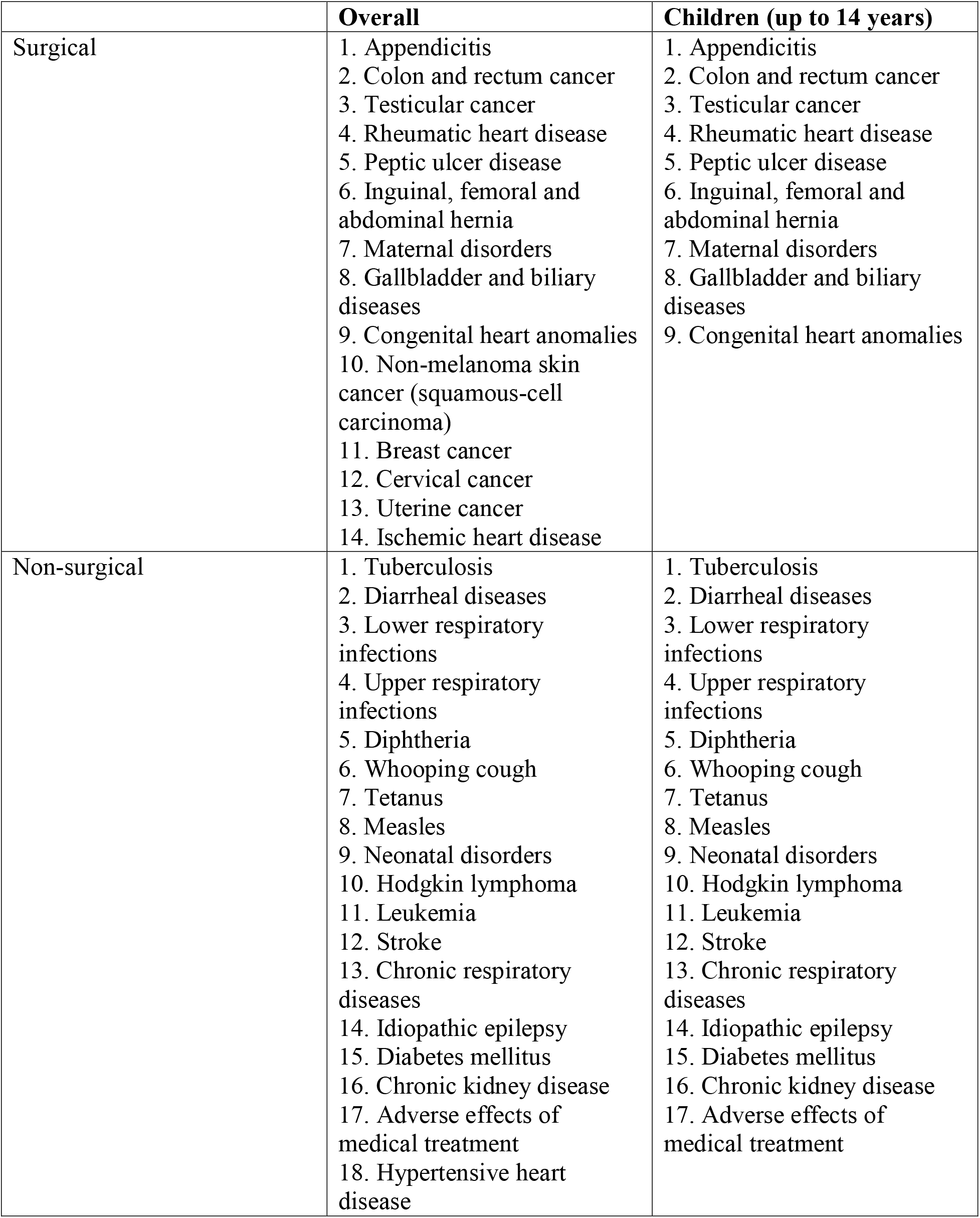
Causes considered for surgical and non-surgical HAQ indices for overall and child populations.

|  | **Overall** | **Children (up to 14 years)** |
| --- | --- | --- |
| Surgical | 1. Appendicitis<br>2. Colon and rectum cancer<br>3. Testicular cancer<br>4. Rheumatic heart disease<br>5. Peptic ulcer disease<br>6. Inguinal, femoral and abdominal hernia<br>7. Maternal disorders<br>8. Gallbladder and biliary diseases<br>9. Congenital heart anomalies<br>10. Non-melanoma skin cancer (squamous-cell carcinoma)<br>11. Breast cancer<br>12. Cervical cancer<br>13. Uterine cancer<br>14. Ischemic heart disease | 1. Appendicitis<br>2. Colon and rectum cancer<br>3. Testicular cancer<br>4. Rheumatic heart disease<br>5. Peptic ulcer disease<br>6. Inguinal, femoral and abdominal hernia<br>7. Maternal disorders<br>8. Gallbladder and biliary diseases<br>9. Congenital heart anomalies |
| Non-surgical | 1. Tuberculosis<br>2. Diarrheal diseases<br>3. Lower respiratory infections<br>4. Upper respiratory infections<br>5. Diphtheria<br>6. Whooping cough<br>7. Tetanus<br>8. Measles<br>9. Neonatal disorders<br>10. Hodgkin lymphoma<br>11. Leukemia<br>12. Stroke<br>13. Chronic respiratory diseases<br>14. Idiopathic epilepsy<br>15. Diabetes mellitus<br>16. Chronic kidney disease<br>17. Adverse effects of medical treatment<br>18. Hypertensive heart disease | 1. Tuberculosis<br>2. Diarrheal diseases<br>3. Lower respiratory infections<br>4. Upper respiratory infections<br>5. Diphtheria<br>6. Whooping cough<br>7. Tetanus<br>8. Measles<br>9. Neonatal disorders<br>10. Hodgkin lymphoma<br>11. Leukemia<br>12. Stroke<br>13. Chronic respiratory diseases<br>14. Idiopathic epilepsy<br>15. Diabetes mellitus<br>16. Chronic kidney disease<br>17. Adverse effects of medical treatment |

A composite index ranging from 0 (worst) to 100 (best) was constructed using an adaptation of the adjusted Mazziotta-Pareto index (AMPI) methodology. AMPI’s advantages include straightforward interpretation, computability, partially compensatory nature, and comparability across times.(10) We constructed HAQ indices for overall and child populations as follows:

1. The indicators, i.e., mortality-to-incidence ratios and risk-standardized death rates for individual causes were normalized by min-max scaling **(Equation 1)**.

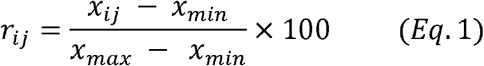

Where *r*_*ij*_ is the normalized indicator for *i*^*th*^ location-year of the 408 country-years and *j*^*th*^ cause of the 14 causes considered for overall surgical, 18 for overall non-surgical, 9 for child surgical, and 17 for child non-surgical HAQ indices.

2. The indicators were aggregated using arithmetic mean, which was then penalized for imbalance using a variance term **(Equations 2)**.

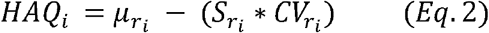

Where HAQ_i_ is the surgical (or non-surgical) index for the *i*^*th*^ location-year, µ is the mean of the normalized indicator values, 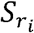 is their standard deviation, and 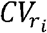 is their coefficient variation.

Post-construction, we conducted the following validation steps for the surgical HAQ indices for overall and child populations. First, Cronbach’s alpha was used to assess internal consistency (α≥0.9: excellent consistency, 0.8≤α<0.9: good, 0.7≤α<0.8: acceptable, and so on). Second, Pearson product-moment correlation was used to measure the association between surgical HAQ indices calculated using the AMPI method and the arithmetic mean approach - followed by the GBD 2019 study to assess the validity of the penalty (variance) term.(2) Third, as basic check whether we observe the expected patterns, nonparametric Kruskal-Wallis rank sum test assessed univariate differences across World Bank income groups. A conservative alpha threshold of 0.01 was used to determine the significance of statistical tests. Finally, as a cursory step toward concurrent validation, we evaluated the Spearman rank correlation between the surgical HAQ indices (for 2019) and the recently described surgical preparedness index (SPI).(11) The SPI assessment included 23 indicators for elective surgical systems and was conducted in 1632 hospitals across 119 countries in 2021. Our analyses included 116 of these that matched the GBD locations.

In aim 2, to track countries’ progress over time, the relative change in the surgical HAQ index was calculated as the ratio of surgical HAQ in 2019 to that in 1990. Values >1 depicted improvement over time. To estimate the average (marginal) values for the World Bank income groups (2019),(12) we used a generalized linear model (GLM) with gamma distribution and log link with the 1990 surgical HAQ as the covariate.

In aim 3, we calculated the country-wise ratio of between the surgical to non-surgical HAQ indices for 2019. Values >1 depicted better surgical access and quality than their non-surgical counterparts. To estimate the marginal values for the World Bank income groups, we used GLM with gamma distribution and log link with 2019 surgical-to-non-surgical HAQ ratio as the outcome and 1990 surgical and non-surgical HAQ index values as the covariates. Analyses and visualization were conducted in R (RStudio Version 1.3.1056).

## Results

### Overall Surgical HAQ Index

In 2019, the overall surgical HAQ varied from 18.00 for the Central African Republic to 98.25 for Canada **(Figure 1a)**. The index depicted excellent internal consistency (Cronbach’s α = 0.94). The index calculated using the AMPI method was significantly and strongly associated with that estimated using the arithmetic mean (n=408 since 204 for 1990 and 2019 each, r=0.99, p<0.0001). The 2019 surgical HAQ index values differed significantly across World Bank income groups (df=3,□^2^=137.1, p<0.0001). The overall surgical HAQ was also significantly associated with the surgical preparedness index (n=116, ρ=0.50, p<0.0001).

**Figure 1:**
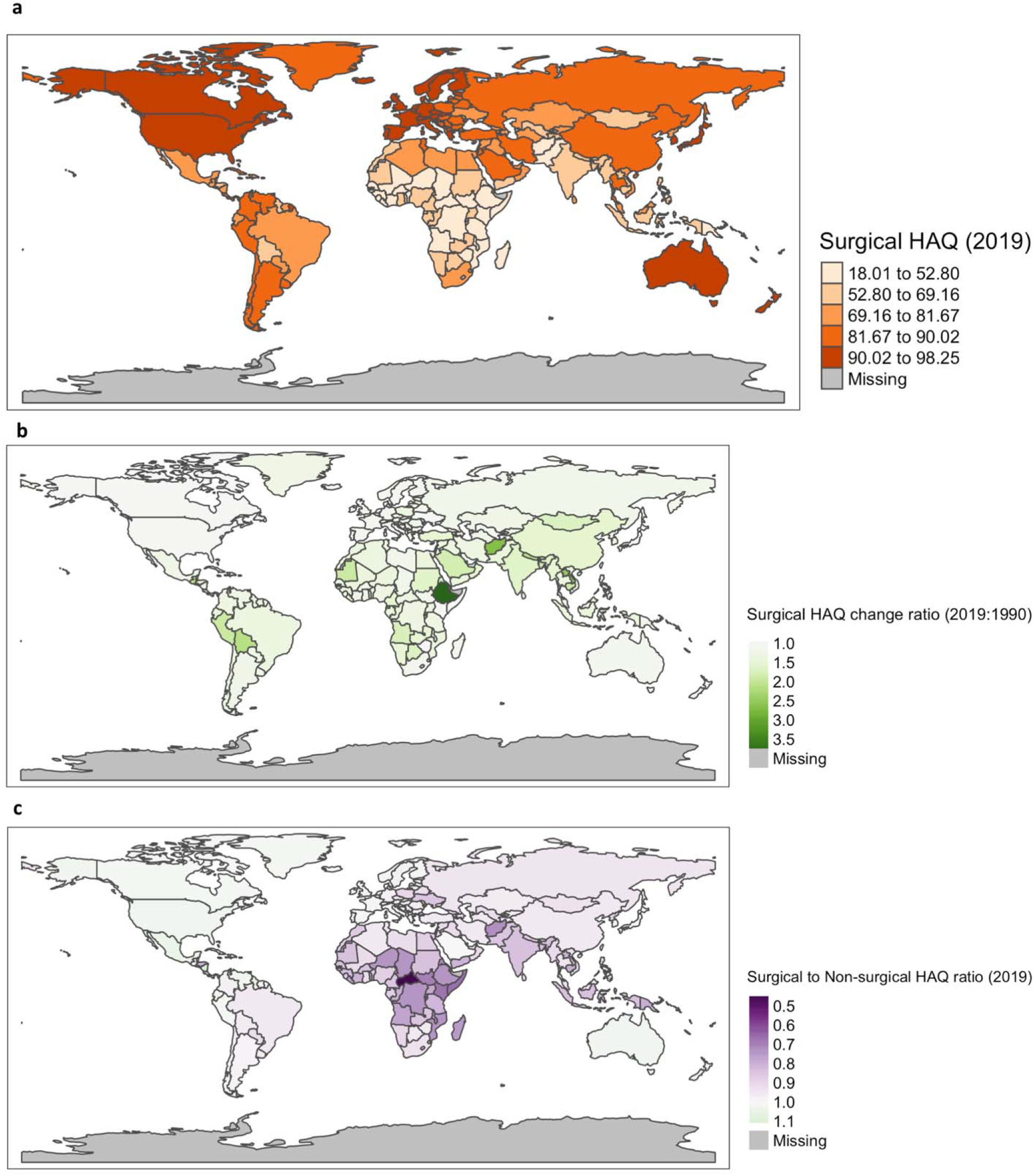
Distribution of a) overall surgical healthcare access and quality (HAQ) index values in 2019, b) raw ratio of 2019 overall surgical HAQ index to 1990 overall surgical HAQ index denoting relative change, and c) raw ratio of 2019 overall surgical to overall non-surgical HAQ indices denoting relative gap across 204 countries and territories. 1a uses a quantile scale while 1b-c use divergent scales centered around the null value (=1).

The overall surgical HAQ index showed the greatest relative change for Ethiopia (2019:1990 ratio=3.88), while Zimbabwe saw a slight worsening (ratio=0.92) **(Figure 1b)**. Except for Zimbabwe and Tajikistan, 202 countries noted progress (>1) in their surgical HAQ from 1990 to 2019. In 2019, Mauritius had the highest overall surgical-to-non-surgical HAQ ratio of 1.16, while surgical HAQ for the Central African Republic lagged the most than its non-surgical HAQ (ratio=0.44) **(Figure 1c)**. Only 31 countries (15.2%) had greater surgical HAQ values than their non-surgical counterparts.

Among the World Bank income groups, after adjusting for the 1990 overall surgical HAQ index, the relative changes in overall surgical HAQ index was greatest for high-income countries, followed by upper-middle-income countries, lower-middle-income, and low-income countries **(Figure 2)**. For all income groups, overall surgical HAQ index values in 2019 were better than in 1990 (adjusted ratios >1). After adjusting for the 1990 overall surgical and non-surgical HAQ indices, the surgical-to-non-surgical ratio in 2019 was greatest for upper-middle-income countries, followed by high-income countries, lower-middle-income countries, and low-income countries **(Figure 2)**. For all income groups, overall surgical HAQ index values were lower than the overall non-surgical HAQ index values, depicting a gap or lag for surgical systems (adjusted ratios <1).

**Figure 2:**
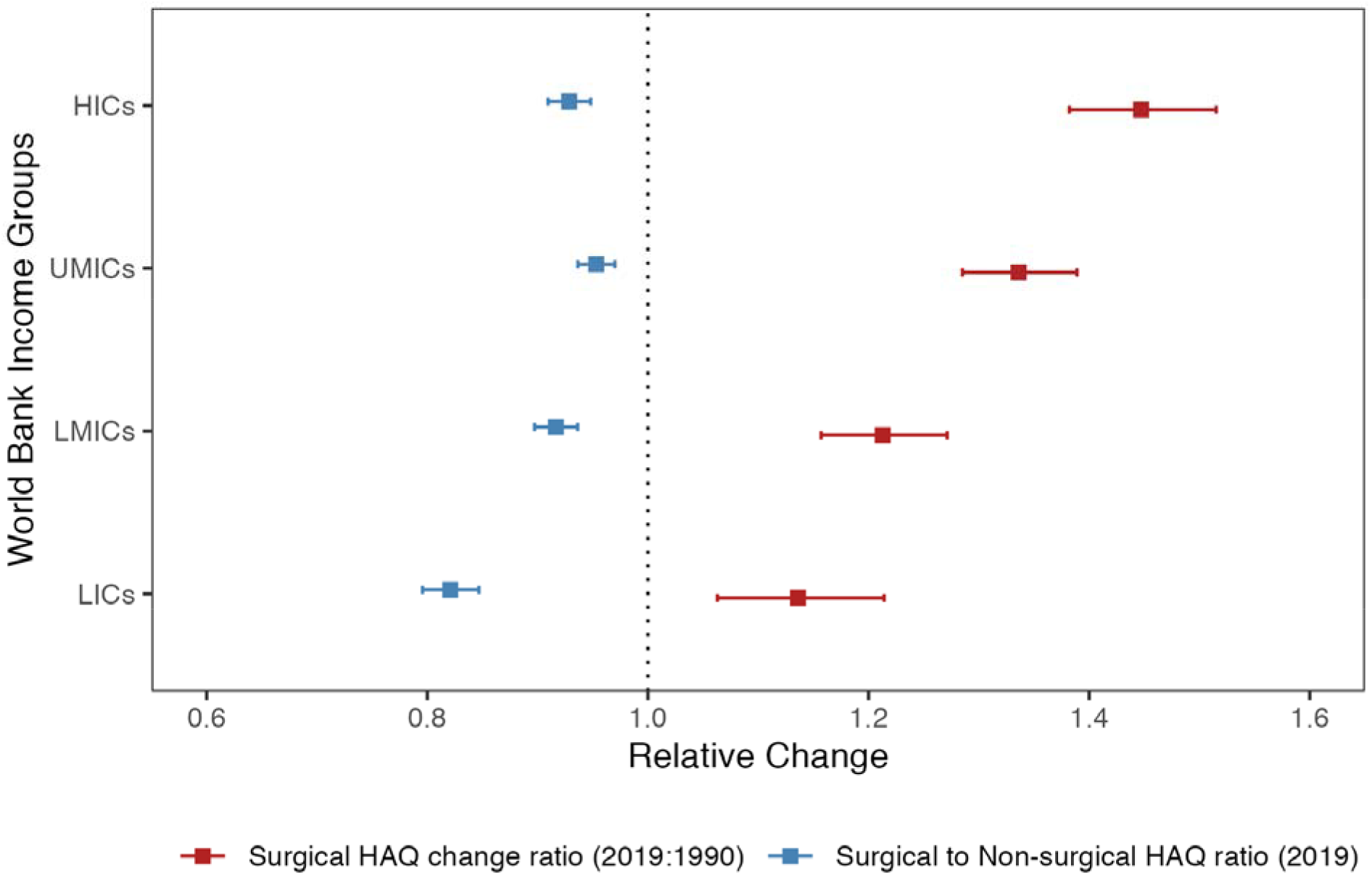
Modeled relative change (2019:1990) ratio for overall surgical HAQ index (red) and relative gap compared to overall non-surgical HAQ index (blue) by World Bank income groups. Both models were gamma regression with log link function. Marginal mean estimates with 95% confidence intervals for the income-group-wise ratios are depicted here. The relative change model (red) was adjusted for the 1990 overall surgical HAQ index value, while the relative gap model was adjusted for the 1990 overall surgical and non-surgical HAQ indices. The dotted vertical line (black) depicts the null value for ratio estimates. HICs = High-income countries, UMICs = Upper-middle-income countries, LMICs = Lower-middle-income countries, LICs = Low-income countries.

### Child Surgical HAQ Index

In 2019, the child surgical HAQ index varied from 39.87 for Chad to 99.41 for San Marino **(Figure 3a)**. The index depicted good internal consistency (Cronbach’s α = 0.86). The index calculated using the AMPI method was significantly and strongly associated with that estimated using the arithmetic mean (n=408 since 204 for 1990 and 2019 each, r=0.99, p<0.0001). The 2019 child surgical HAQ index values differed significantly across World Bank income groups (df=3, □^2^=133.67, p<0.0001). The child surgical HAQ index was also significantly associated with the surgical preparedness index (n=116, ρ=0.51, p<0.0001).

**Figure 3:**
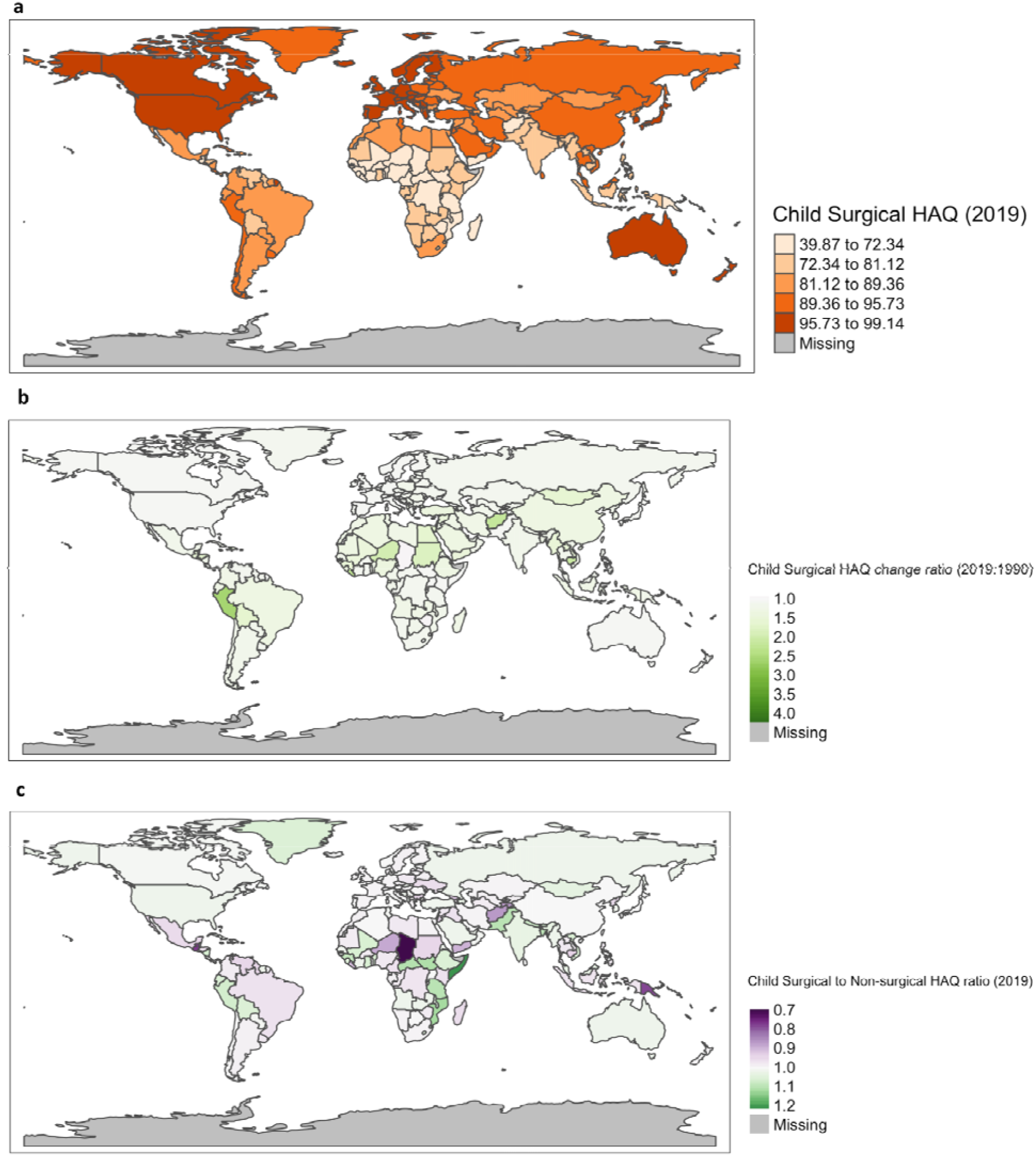
Distribution of a) child surgical healthcare access and quality (HAQ) index for children in 2019, b) raw ratio of 2019 to 1990 child surgical HAQ indices denoting relative change, and c) raw ratio of 2019 child surgical to non-surgical HAQ index denoting relative gap across 204 countries and territories. 1a uses a quantile scale while 1b-c use divergent scales centered around the null value (=1).

The child surgical HAQ index showed the largest relative improvement for Haiti (ratio=4.41) from 1990 to 2019, while Chad saw a small worsening (ratio=0.94) **(Figure 3b)**. Except Zimbabwe and Chad, others saw progress (values >1) in child surgical HAQ index from 1990 to 2019. The relative gap compared to nonsurgical HAQ ranged from 0.38 for Liberia to 2.12 for Haiti in 2019 **(Figure 3c)**. In 61.28% of countries, the child surgical HAQ index lagged behind non-surgical HAQ index.

Among the World Bank income groups, after adjusting for the 1990 child surgical HAQ index, the relative changes in the child surgical HAQ index was greatest for high-income countries, followed by upper-middle-income countries, lower-middle-income, and low-income countries **(Figure 4)**. For all income groups, overall surgical HAQ index values in 2019 were better than in 1990 (adjusted ratios >1). After adjusting for the 1990 child surgical and non-surgical HAQ indices, the surgical-to-non-surgical ratio in 2019 was greatest for lower-middle-income countries, followed by upper-middle-income countries, high-income countries, and low-income countries **(Figure 4)**. However, the differences across World Bank income groups were less pronounced than those overserved for overall surgical HAQ index. For all income groups, child surgical HAQ index values were lower than the non-surgical counterparts, depicting a gap or lag for surgical systems (adjusted ratios <1. However, the 95% confidence intervals crossed the null value of 1 for all groups.

**Figure 4:**
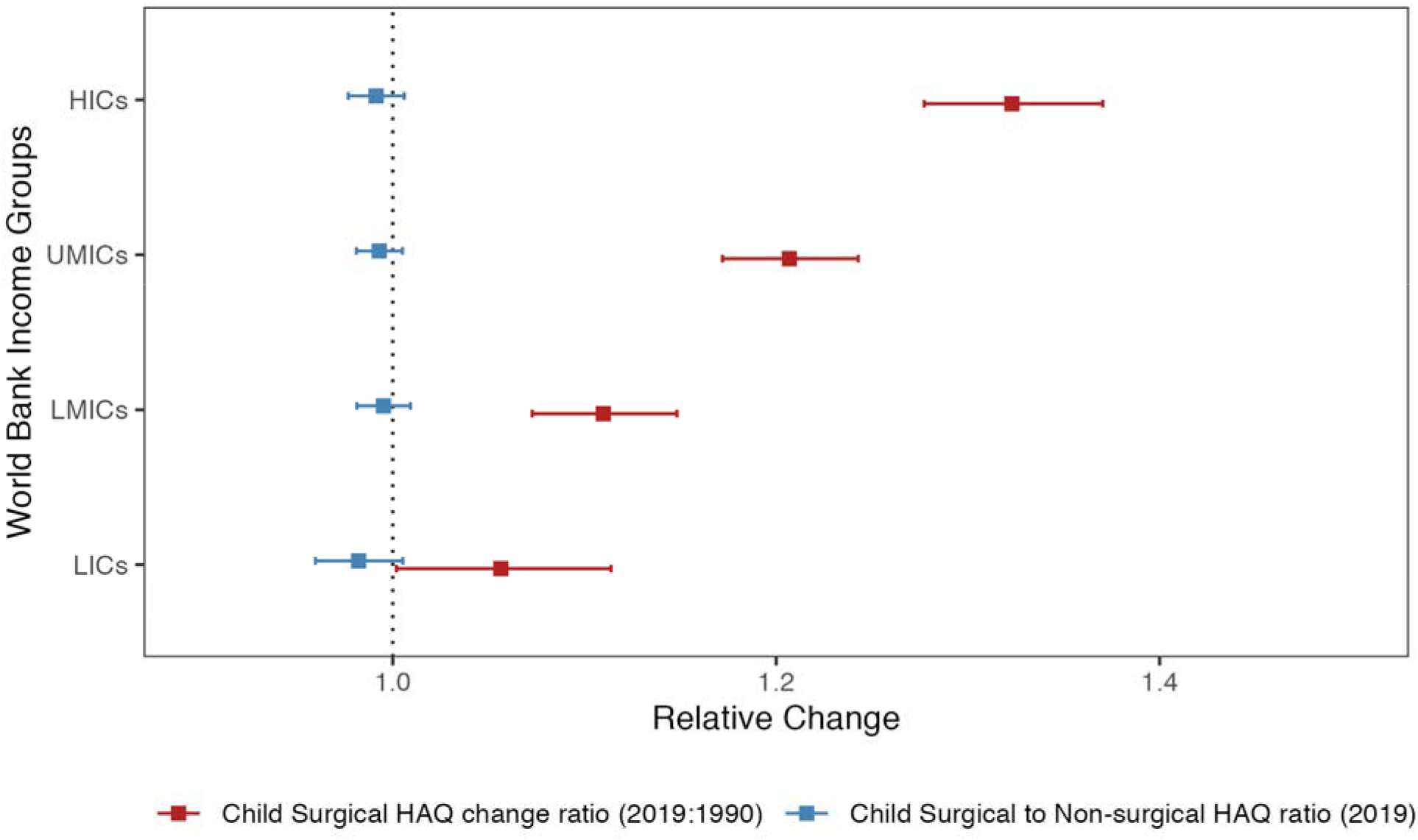
Modeled relative change (2019:1990) ratio for child surgical HAQ index (red) and relative gap compared to child non-surgical HAQ index (blue) by World Bank income groups. Both models were gamma regression with log link function. Marginal mean estimates with 95% confidence intervals for the income-group-wise ratios are depicted here. The relative change model (red) was adjusted for the 1990 child surgical HAQ index, while the relative gap model was adjusted for the 1990 child surgical and non-surgical HAQ indices. The dotted vertical line (black) depicts the null value for ratio estimates. HICs = High-income countries, UMICs = Upper-middle-income countries, LMICs = Lower-middle-income countries, LICs = Low-income countries.

## Discussion

### Summary and Interpretation

The current study presents an up-to-date global analysis indexing surgical healthcare access and quality index for 204 countries and territories. We found that low-income countries, mostly in sub-Saharan Africa, had the lowest overall and child surgical HAQ values while high- and upper-middle -income countries had the highest index values. For World Bank income groups, the patterns of progress over time were similar for overall and child surgical HAQ indices from 1990 to 2019. The overall and child populations across income groups differed for the relative gap between surgical and non-surgical HAQ indices. We found that low-income countries had limited progress in surgical HAQ indices since 1990 and lagged behind non-surgical counterparts in 2019.

Surgical care is integral to universal health coverage (UHC). More specifically, within UHC, capacity to provide surgery to all is a marker of robust secondary and tertiary levels of care in a health system.(13, 14) Over the last thirty years, building public system’s capacity for secondary and higher levels of care has suffered neglect globally.(15, 16) Neglect toward surgical care systems is noted in health policies and financing.(17–20) This neglect is reflected in the limited progress for surgical HAQ index and the relative lag compared to the non-surgical HAQ. Improvements in surgical care can have positive spillover effects on wider care systems since the underlying infrastructure and workforce is shared across clinical specialties. Hence, investing in surgical care delivery is a major component of broader health system strengthening. Further, epidemiological transition from infectious disease burden to noncommunicable disease and injuries requires centering surgical care. For instance, the under-5 mortality rates across low- and middle-income countries have dropped in the recent decades, predominantly due to primary care interventions. However, the share of deaths due to congenital conditions requiring surgical interventions has increased – necessitating investing in children’s surgery known to have good return on investment.(21–23)

### Research and Policy Implications

Our findings are relevant for international funders and intergovernmental and national policymakers in the following ways:

a. For low-income countries, the limited improvement in surgical HAQ from 1990 to 2019 and the current gap compared to the non-surgical HAQ necessitates financial investments and policy attention for surgical system strengthening. Funding agencies and philanthropies can use the index for allocating resources. For instance, countries with a greater burden of surgically amenable mortality, i.e., those in the lowest quartile of surgical HAQ index values could be prioritized for funding. Those with a track record of progress could also be prioritized for developmental loans to ensure continued trends.
b. Pediatric surgery needs to be prioritized in low- and lower-middle-income countries. Multiple studies have noted limited attention to surgical care of children in the national health policies, strategies, and plans (NHPSPs) as well as the national surgical, obstetric, and anesthesia plans (NSOAPs).(6, 20, 24, 25) Such lack of policy attention is stark given that the majority of world’s children live in these countries. The gap in the child surgical HAQ relative to the non-surgical counterpart and limited progress since 1990 could potentially be explained by the limited policy attention and financing. Including pediatric surgery in existing health programs for children could be one way of ensuring pediatric surgery prioritization.(23, 26) For instance, previously we have noted that increasing trained pediatric surgical workforce and including more packages for children’s surgery in government-funded health insurance schemes could be vital for developing economies.(21, 27)
c. Integrating the surgical HAQ index into health monitoring and evaluation systems would help measure the population health impacts of policy and programmatic interventions, including NSOAPs.(28) The LCoGS indicators look at preparedness, delivery and financial impact of the surgical care.(29) Changes in these indicators in response to implementing an NSOAP could be only perceived as changes in the process outcomes. However, true impact can only be measured by assessing population health outcomes enabled by composite measures such as the surgical HAQ index.
d. It can also be used for benchmarking the minimum resources, including the surgical workforce needed. For instance, previous studies including have used outcomes such as the maternal mortality ratio and infant mortality rate to determine the minimum number of required surgeons, anesthetists, and obstetricians.(27, 30) However, such unidimensional measures may not capture the wide array of health outcomes that need surgical workforce. A composite such as the surgical HAQ index can help circumvent the problem.
e. The current findings can help assist Global Surgery advocacy in multiple ways. The evidence on the gap between surgical and non-surgical HAQ indices is useful to advocate for the need to prioritize surgical care across various ongoing initiatives including those for pediatric surgery, primary healthcare care, and emergency, critical and operative care. The progress made by surgical care systems over the last decades can be used by Global Surgery advocates to inspire confidence and provide a positive narrative for continued investments for surgical care in low- and middle-income countries.

### Strengths and Limitations

The surgical HAQ index computed using an adaptation of the adjusted Mazziotta-Pareto index methodology had good internal consistency, was correlated strongly with the arithmetic mean, and had a statistically significant correlation with the surgical preparedness index, denoting concurrent validity. Surgical HAQ constructed here shares the theoretical framework with past attempts.(4) However, the computations vary in important ways. We used the AMPI methodology that produces a partially compensatory index. This means that poor performance on one indicator cannot be compensated by progress on another, which is an advantage over using an arithmetic mean. From the perspective of policymaking and resource allocation, all aspects of preventable mortality are prioritized adequately. The AMPI-derived HAQ is easily interpretable and stable compared to principal component analysis (PCA). The GBD also shifted from calculating HAQ using PCA to using arithmetic mean.(1, 2) Our approach ensures that the index is reliably replicable in future assessments and is not data-dependent.

Since our analysis relies on the modeled estimates from IHME, it inherits the limitations of the GBD estimates including the lack of considerations for within-country equity or cultural acceptability of certain healthcare interventions and a possibility of misclassification of causes of mortality in the original GBD modeling process. Second, we lack uncertainty estimates that should be corrected in the future. Third, the classification for surgical vs. non-surgical causes is based on the previous coarse definition. However, more granular and consensus-driven classifications are needed. Fourth, the current study could not investigate the impact of the COVID-19 pandemic on surgical healthcare access and quality. However, the next iteration of the Global Burden Disease study (GBD 2021) should permit that. Lastly, the surgical conditions for children that were present in the HAQ cause list do not include several common pediatric surgical conditions. Therefore, our results for children’s surgery may deviate from the actual progress or current need in some low- and middle-income countries. Regardless, we provide updated pre-pandemic estimates for the surgical and non-surgical HAQ indices that provide a baseline assessment useful for policymaking across countries.

## Conclusions

The differences across countries, notable progress over the last three decades, and gap relative to non-surgical HAQ depict global disparities in preventable mortality due to access to quality surgical care. Surgical HAQ index is a composite with important implications for future research and policymaking. Closing the gap between the surgical and non-surgical HAQ indices is not only crucial for the surgical care but also for improving overall healthcare given surgery’s inevitable role in the health system strengthening and universal health coverage.

## Data Availability

All data produced in the present study are available upon reasonable request to the authors.

## Abbreviations

HAQ: Healthcare Access and Quality
GBD: Global Burden of Disease
AMPI: Adjusted Mazziotta-Pareto Index
HICs: High-income countries
UMICs: Upper-middle-income countries
LMICs: Lower-middle-income countries
LICs: Low-income countries
NSOAP: National surgical, obstetric, and anesthesia plan
NHPSP: National health policy, strategy, and plan
UHC: Universal health coverage

## Declarations

### Author Contributions

**<u>Siddhesh Zadey:</u>** Conceptualization, Methodology, Formal Analysis, Data Curation, Writing - Original Draft, Writing - Review & Editing, Project Administration.

**<u>Emily R.Smith:</u>** Writing - Review & Editing.

**<u>Catherine A.Staton:</u>** Funding Acquisition, Writing - Review & Editing.

**<u>Tamara N.Fitzgerald:</u>** Funding Acquisition, Writing - Review & Editing.

**<u>Joao Ricardo Nickenig Vissoci:</u>** Funding Acquisition, Supervision, Writing - Review & Editing.

## Acknowledgments

None

## Funding

Global Health Equity Pilot Award, Duke Global Health Institute

## Competing interests

Siddhesh Zadey is the co-founding directors of the Association for Socially Applicable Research (ASAR). He also represents ASAR at the G4 Alliance Permanent Council Member and serves as the Chair of the SOTA Care in Asia Working Group, The G4 Alliance. Other authors declare no conflicts.

## Ethics approval and consent to participate

Not applicable since the study involves analysis of publicly available aggregate data.

## Consent for publication

Not applicable.

## Availability of data and materials

Data used and produced in the manuscript are available upon reasonable request from the authors.

## Notes

### Funding Statement

This study was funded by Global Health Equity Pilot Award, Duke Global Health Institute.

### Summary of Updates

We have added a full analysis for child surgical HAQ index. Significant changes have been made to the manuscript's title, abstract, results and discussion sections. Additionally, updates have been made to funding and disclosures based on the authors' current status.

